# Central and peripheral nervous system complications of COVID-19: A prospective tertiary center cohort with 3-month follow-up

**DOI:** 10.1101/2020.11.15.20231001

**Authors:** Vardan Nersesjan, Moshgan Amiri, Anne-Mette Lebech, Casper Roed, Helene Mens, Lene Russel, Lise Fonsmark, Marianne Berntsen, Sigurdur Thor Sigurdsson, Jonathan Carlsen, Annika Langkilde, Pernille Martens, Eva Løbner Lund, Klaus Hansen, Bo Jespersen, Marie Norsker Folke, Per Meden, Anne-Mette Hejl, Christian Wamberg, Michael E. Benros, Daniel Kondziella

**Author notes:** **Correspondence:** Daniel Kondziella, MD, MSc, PhD, FEBN, and Michael E. Benros, MD, PhD. **Conflicts of interest:** None.

## Abstract

**Objective:** To systematically describe CNS and PNS complications in hospitalized COVID-19 patients.

**Methods:** We conducted a prospective, consecutive, observational study of adult patients from a tertiary referral center with confirmed COVID-19. All patients were screened daily for neurological and neuropsychiatric symptoms during admission, at discharge and at 3-month follow-up. We classified complications as caused by SARS-CoV-2 neurotropism, immune-mediated or critical illness-related.

**Results:** From April-September 2020, we enrolled 61 consecutively admitted COVID-19 patients, 35 (57%) of whom were referred to ICU for respiratory failure. Evaluation revealed a higher frequency of CNS/PNS symptoms in ICU patients compared to non-ICU patients. The most common CNS complication was encephalopathy (n=22, 36.1%), which was severe in 13 patients (GCS≤12), including 8 with akinetic mutism. Length of ICU admission was an independent predictor of encephalopathy (OR=1.23). Other CNS complications included ischemic stroke, a biopsy-proven acute necrotizing encephalitis, and transverse myelitis. The most common PNS complication was critical illness polyneuromyopathy (13.1%), with prolonged ICU stay as independent predictor (OR=1.14). Treatment-related PNS complications included meralgia paresthetica. Of 41 complications in total, 3 were classified as para/post-infectious. The remainder included cases secondary to critical illness or other causes (n=34) or without sufficient investigations (n=4). Cerebrospinal fluid was negative for SARS-CoV-2 RNA in all 5 patients investigated.

**Conclusions:** CNS/PNS complications were common in hospitalized COVID-19 patients, particularly in ICU patients, and often attributable to critical illness. In cases with COVID-19 as the primary cause for neurological disease, there were no signs of viral neurotropism, but laboratory changes suggested autoimmune-mediated mechanisms.

## 1. Introduction

Central (CNS) and peripheral (PNS) nervous system manifestations can occur during and after coronavirus disease 2019 (COVID-19) [1–6], but the underlying mechanisms, the semiology and the frequency of these complications remain poorly understood [7,8].

Limited postmortem studies have shown signs of hypoxic brain injury [9] and neuro-inflammatory changes in the brainstem [10], while neuropathological data from the PNS are so far almost non-existent. As to the etiology, direct invasion of severe acute respiratory syndrome coronavirus 2 (SARS-CoV-2) into neuronal tissue has been suggested in a few cases [11,12], but autoimmune-mediated injury and neurological complications related to intensive care management are also recognized [7,8]. The contribution of these mechanisms to the overall burden of COVID-19 CNS and PNS complications is unknown. Furthermore, the reported prevalence of these complications varies widely, from 3.5 to 84% [1–6], owing to the heterogeneity in methodology and case definitions. Some reports include descriptions of neurological symptoms but no confirmatory laboratory investigations, which makes the distinction between primary COVID-19-and secondary critical illness-induced complications difficult [5]. For instance, more than one half of patients with sepsis show signs of sepsis-associated encephalopathy [13] and a further 70% of those with sepsis-associated encephalopathy develop critical illness polyneuromyopathy (CIPM) [14]. Also, many studies report on observations made during neurological consultation, thereby missing cases that are not referred to a neurologist [15]. Finally, most reports include convenience samples, lack follow-up and do not attempt to determine if COVID-19 complications are caused by direct viral invasion of the nervous system, by para/post-infectious immune-mediated mechanisms or by treatment-related injury [7].

Here, we conducted a prospective observational study of a complete and consecutive sample of COVID-19 patients from a tertiary referral center, including 3-month follow-up. Our aim was to describe the frequency and phenomenology of CNS and PNS complications, as well as their presumed origin as determined by neurological work-up, during and after SARS-CoV-2 infection. The primary endpoints were the number, clinical phenotypes and mechanisms in patients with COVID-19-related CNS and PNS complications compared to all patients admitted with COVID-19. We hypothesized that these complications would be more frequent and serious in the most severely affected COVID-19 patients and that immune-mediated and treatment-related complications would be more prevalent than those related to SARS-CoV-2 neurotropism.

## 2. Methods

### 2.1 Study population

From April to September 2020, we prospectively enrolled SARS-CoV-2 positive patients ≥18 years of age, admitted to Rigshospitalet, Copenhagen University Hospital, a tertiary referral center in Copenhagen, Denmark. Patients were enrolled from the hospital’s newly established COVID-19 intensive care unit (COVITA), the COVID-19 intermediate care unit (COVIMA) and the neurological department. Two of the authors (VN, MA), supervised by a board-certified neurologist with experience in neurocritical care (DK), screened all three departments each morning for newly admitted patients with COVID-19. All included patients had positive SARS-CoV-2 polymerase-chain reaction (PCR) by nasopharyngeal/tracheal testing, except for 1 patient in whom a clinical suspicion of COVID-19 was confirmed 27 days after symptom onset by a strongly positive SARS-CoV-2 IgG antibody titer. We followed all patients prospectively from the time of enrollment to discharge and 3-month follow-up or death. Patients were prospectively screened for neurological and psychiatric symptoms during admissions at COVITA and COVIMA and evaluated for cognitive status and neurological deficits at discharge. Patients enrolled from the neurological department were screened for suspected neurological complications during/after a COVID-19 infection.

### 2.2 Clinical screening during admission

We used a prespecified screening tool made to detect neurological and psychiatric symptoms (see appendix, **Figure S1**) during admission at COVITA and COVIMA. The screening tool was incorporated as part of the daily clinical rounds by the attending physicians, and patients were examined systematically at the bedside to detect CNS and PNS symptoms and/or deficits. Each day, all COVID-19 patients were thus screened for the following:

I. Consciousness level as assessed by Glasgow Coma Scale (GCS) and Full-Outline of UnResponsiveness (FOUR) scores. When appropriate, sedated ICU patients were assessed following a wake-up call and categorized as showing either ‘rapid recovery’, ‘slow awakening’ (defined as prolonged recovery following >24h clinical unresponsiveness after stop of sedation) and ‘no awakening’ (defined as coma or vegetative state/unresponsive wakefulness syndrome persisting for ≥24h).
II. Complaints of headache and somatosensory symptoms, i.e. anosmia and/or ageusia, pain, and paresthesia.
III. Cognitive deficits, motor palsies, sensory deficits, cranial nerve deficits and cerebellar affection (e.g. ataxia or nystagmus) as determined by clinical neurological exam.
IV. Seizures, irrespective of supposed origin (i.e. epileptic versus non-epileptic).
V. Presence of delirium, defined as presence of auditory or visual hallucinations, delusional thoughts such as paranoia, psychomotor agitation or reduction and/or fluctuating consciousness with disorientation.
VI. Emotional lability defined as exaggerated or uncontrollable laughing, crying or anger disproportionate to the circumstances.

Further, we collected routine clinical and laboratory data including age, sex, medication, previous medical history, and results from brain imaging (CT or MRI) during admission, cerebrospinal fluid (CSF) results, electroencephalography (EEG) findings, nerve conductions studies (ENG) and electromyography (EMG), as well as modified Rankin Scale (mRS) prior to admission, at discharge and at 3-month follow-up.

Two board-certified neuroradiologists systematically re-evaluated all clinical neuroimaging (if present) of all enrolled patients during admission and after discharge, including CT, CT angiography and MRI, according to a prespecified protocol, classifying burden, localization, and (possible) etiology of acute and chronic (i.e. unrelated to the present admission) pathologies.

### 2.3 Discharge evaluation

At discharge (i.e. 0-4 days prior to discharge), we evaluated patients with a full neurological exam, including the Montreal Cognitive Assessment (MOCA). When a full MOCA was not possible (e.g. owing to the patient’s lack of English or Danish language proficiency), we examined with a clock-drawing test to assess executive function and search for hemi-spatial neglect, and a simple 5 minutes 3-word recall test to assess memory function. Gait function was categorized as normal, assisted or bedridden/wheelchair.

### 2.4 Three-month follow-up

Enrolled patients were followed up 3 months after discharge via electronic health records (EHR). We obtained follow-up data on mRS, re-admissions (if any), death after discharge, and new-onset neurological and psychiatric diagnoses. If mRS data was not available from the EHR, we obtained the data via telephone interview with the patient or next-of-kin.

### 2.5 Prespecified complications

Based on our observed findings from 2.2 to 2.4, we screened for the following prespecified CNS and PNS complications occurring during admission or after discharge and 3-month follow-up:

CNS: a) *stroke* (i.e. ischemic or hemorrhagic, or subarachnoid hemorrhage; b) *encephalopathy* [16], i.e. change in personality, behavior, cognition or consciousness, including delirium; and categorized as “mild”, when behavioral and/or cognitive changes occurred without reduced consciousness (GCS≥13), or “severe” (all other), and reduced consciousness was only noted when patients were off sedation; c) *encephalitis* [17], i.e. encephalopathy PLUS new focal CNS deficits and/or new-onset epileptic seizures PLUS CSF inflammation (e.g. pleocytosis) and/or MRI changes and/or brain pathology suggesting brain inflammation; d) *myelitis*, i.e. clinical signs of medullary affection and MRI confirmed changes suggesting inflammatory lesions; and e) *primary psychiatric disorders*, including depression, anxiety, post-traumatic stress disorder, obsessive compulsive disorder or psychotic disorder diagnosed by ICD-10 criteria.

PNS: a) *neuropathies*, including mono-(e.g. peripheral facial palsy) and polyneuropathies (e.g. Guillain Barré syndrome); b) *acute myopathies*, defined as myalgia with motor paresis and biochemical evidence of muscle degradation (e.g. elevated creatine kinase or myoglobin at least 5 times the upper limit of normal) [18]; and c) *CIPM*, i.e. defined as limb weakness with clinical signs of lower sensory and motor neuron deficits following sepsis, multi-organ failure, respiratory failure, and/or septic inflammatory response syndrome; categorized as ‘definite’ with consistent ENG/EMG abnormalities or ‘possible’ without neurophysiological exam.

### 2.6 Case definitions of SARS-CoV-2-associated neurological disease

Based on the recently proposed case definition of neurological complications owing to COVID-19,[10] we assigned the observed complications (2.5) to one of the following disease mechanisms:

A. *Direct viral invasion*. (1) Evidence of SARS-CoV-2 detected in CSF or evidence of SARS-CoV-2-specific intrathecal antibody production; and (2) no other explanatory pathogen or cause found.
B. *Immune-mediated mechanisms*. (1) Neurological disease onset within 6 weeks of acute infection; (2) no evidence of other commonly associated causes (e.g. recently or concomitant infection with campylobacter, cytomegalovirus (CMV), Epstein-Barr virus (EBV), herpes simplex (HSV) or varicella virus (VZV)) and (3) paraclinical evidence of immune-mediated mechanisms such as inflammatory lesions on MRI and/or CSF pleocytosis or oligoclonal bands and/or brain pathology findings.
C. *Complications secondary to critical illness*, management-related or other causes. (1) Other, more likely causes such as delirium, hypoxia, sepsis, metabolic derangement or other complications to critical illness (e.g. septic or hypoxic encephalopathy) and (2) if investigated with brain imaging, EEG or CSF study, there are no signs of disease mechanism A or B.
D. *Insufficiently investigated*. (1) No explainable cause found, therefore not categorized as C, and (2) not investigated with relevant CSF study and/or neuroimaging to confirm or dismiss A or B disease mechanisms.

Patients were classified in the case definitions by two observers (VN, MA) with >4 years neurological experience based on consensus. Disagreement was settled by a third observer (DK) with >15 years of neurological experience.

### 2.6 Statistical analysis

Based on the data from clinical screening during admission (2.2) and at 3-month follow-up (2.4), we assessed if patients had developed CNS or PNS complications and categorized these complications as having occurred ‘during admission’ or ‘after admission’, i.e. patients who were not admitted for an active SARS-CoV2 infection but admitted for a neurological complication following COVID-19 were categorized as ‘after admission’. Patients were dichotomized in ICU and non-ICU admission and demographic variables, neurological and neuropsychiatric symptoms during admission and discharge, paraclinical findings and 3-month follow-up characteristics were compared using the student’s t test for normally distributed values, the Mann-Whitney U test (Wilcoxon rank-sum) for non-normally distributed continuous variables and Fisher’s exact test for categorical values. Variables for binary logistic regression for the development of severe encephalopathy or CIPM were selected based on biological plausibility and bivariate associations seen in our data. A 2-sided p<0.05 was considered significant. We did not correct for multiple testing. All analyses were performed using Stata/IC 15.1 (StataCorp. 2017, College Station, TX).

### 2.7 Ethics statement

The Ethics Committee of the Capital Region of Denmark approved the study and waived the need for written consent because the risks were deemed negligible (reference j.nr. 20025838).

## 3. Results

From April 1 to September 25, 2020, we prospectively enrolled 61 COVID-19 patients ≥ 18 years, consecutively admitted to Rigshospitalet, Copenhagen University Hospital. Mean age was 62.7 years, and the male-to-female ratio was 1.7:1 (63% males). Prior to COVID19, somatic comorbidities were known in 52 (85%), neurological comorbidities in 11 (18%) and psychiatric comorbidities in 4 (6.6%) of the 61 patients. Mean days from COVID-19 symptom onset to hospital admission was 7.2, and mean admission duration was 29.7 days. Seventeen patients (27.9%) were treated with renal replacement therapy and 35 patients (57.4%) were admitted to the ICU, of which 34 (97%) were mechanically ventilated (21 in the prone position, 61.7%) and 6 (17.1%) treated with extracorporeal membrane oxygenation (ECMO). Ten (16.4%) patients died in-hospital, and 3-month follow-up was available for 45 patients. See **Table 1** for full outline of basic demographics including previous medical history and admission characteristics, and **Figure 1** for an overview over patient enrollment and observed complications. In the following, results are listed according to symptoms (**Tables 2-4**), investigations (**Figure 2** and **Table 5**) and clinical diagnoses (**Table 6**).

**Table 1.**
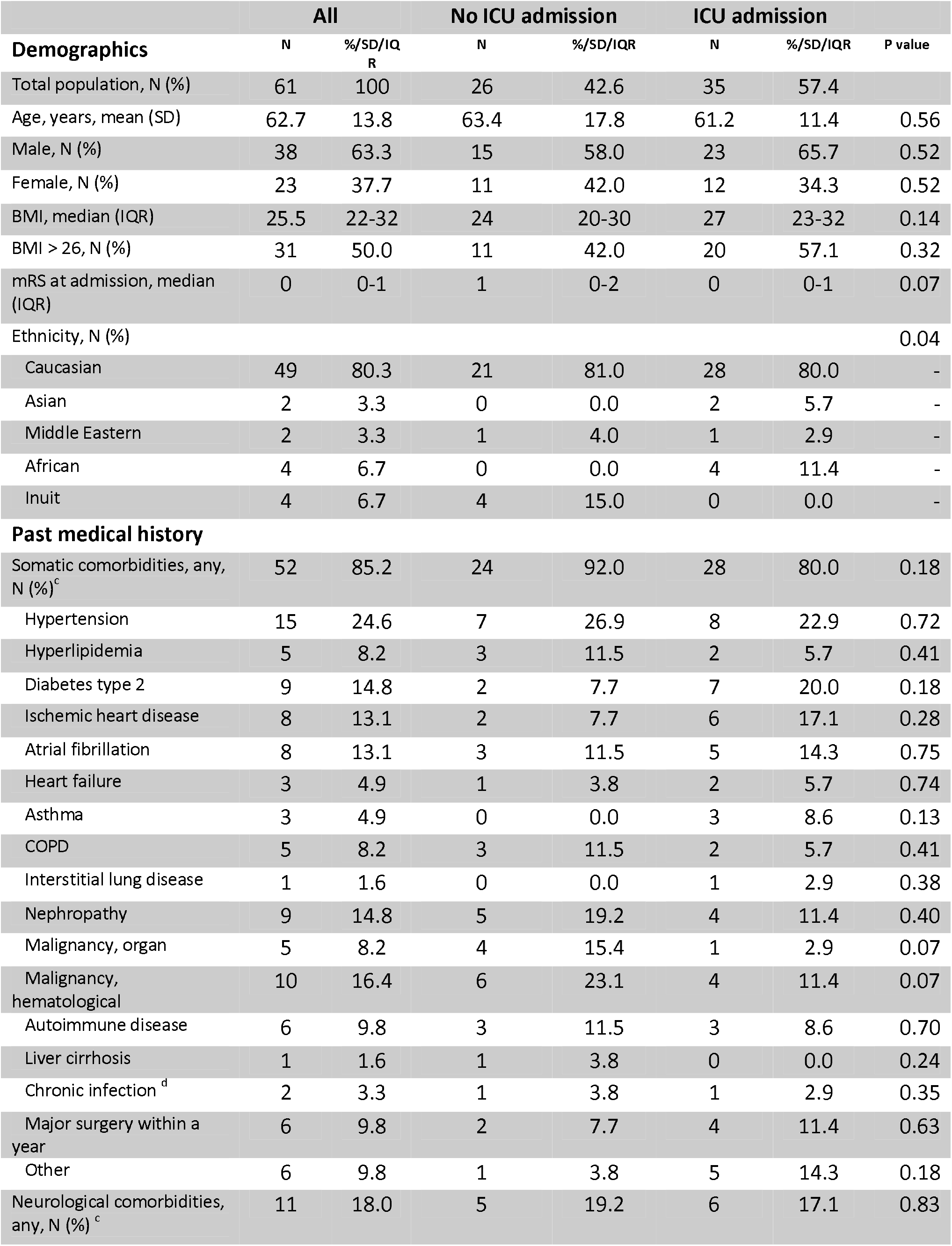
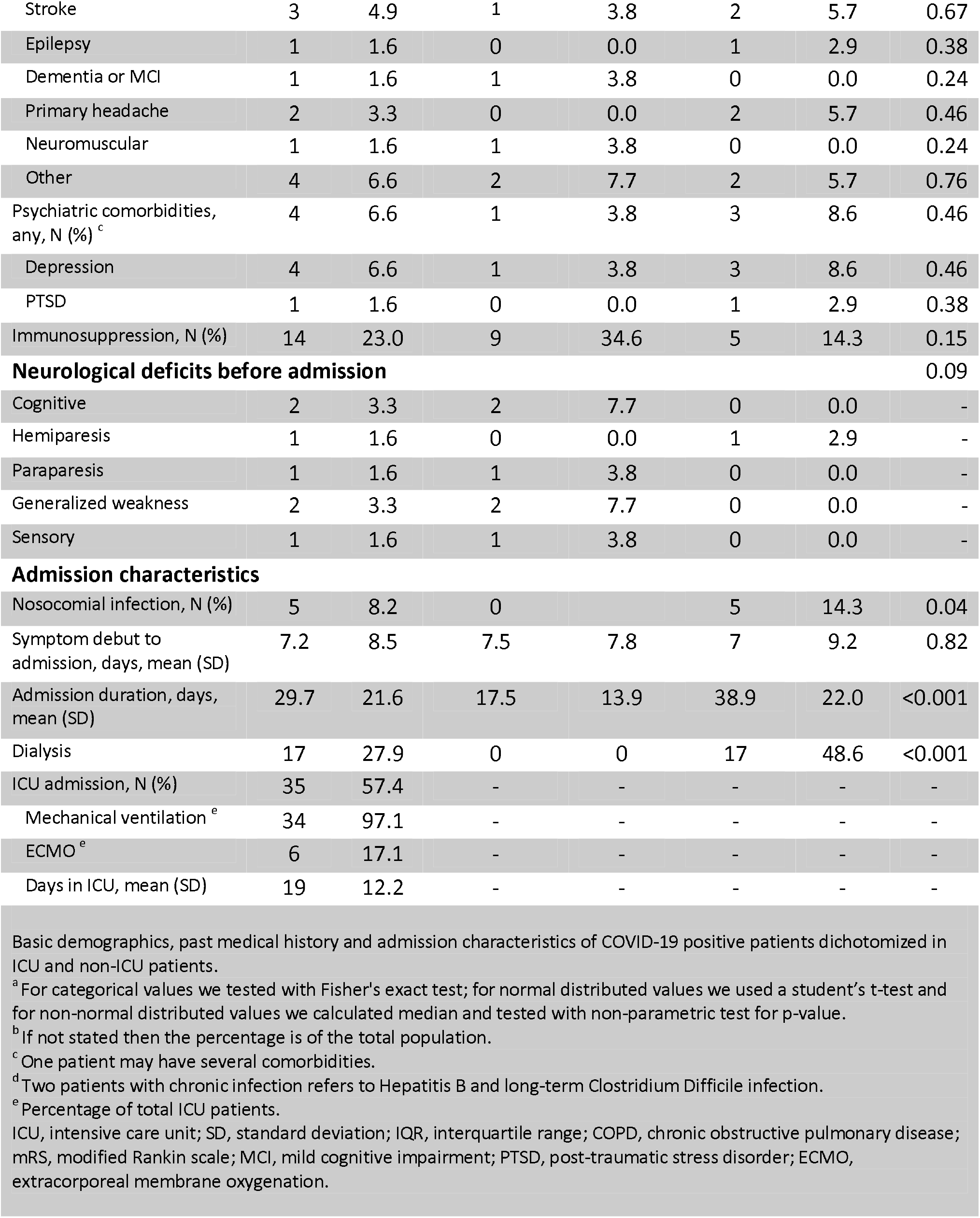
Demographics and admission characteristics ^a,b^.

|  | All |  | No ICU admission |  | ICU admission |  |  |
| --- | --- | --- | --- | --- | --- | --- | --- |
| **Demographics** | N | %/SD/IQR | N | %/SD/IQR | N | %/SD/IQR | P value |
| Total population, N (%) | 61 | 100 | 26 | 42.6 | 35 | 57.4 |  |
| Age, years, mean (SD) | 62.7 | 13.8 | 63.4 | 17.8 | 61.2 | 11.4 | 0.56 |
| Male, N (%) | 38 | 63.3 | 15 | 58.0 | 23 | 65.7 | 0.52 |
| Female, N (%) | 23 | 37.7 | 11 | 42.0 | 12 | 34.3 | 0.52 |
| BMI, median (IQR) | 25.5 | 22-32 | 24 | 20-30 | 27 | 23-32 | 0.14 |
| BMI > 26, N (%) | 31 | 50.0 | 11 | 42.0 | 20 | 57.1 | 0.32 |
| mRS at admission, median (IQR) | 0 | 0-1 | 1 | 0-2 | 0 | 0-1 | 0.07 |
| Ethnicity, N (%) |  |  |  |  |  |  | 0.04 |
| Caucasian | 49 | 80.3 | 21 | 81.0 | 28 | 80.0 | - |
| Asian | 2 | 3.3 | 0 | 0.0 | 2 | 5.7 | - |
| Middle Eastern | 2 | 3.3 | 1 | 4.0 | 1 | 2.9 | - |
| African | 4 | 6.7 | 0 | 0.0 | 4 | 11.4 | - |
| Inuit | 4 | 6.7 | 4 | 15.0 | 0 | 0.0 | - |
| **Past medical history** |  |  |  |  |  |  |  |
| Somatic comorbidities, any, N (%)<sup>c</sup> | 52 | 85.2 | 24 | 92.0 | 28 | 80.0 | 0.18 |
| Hypertension | 15 | 24.6 | 7 | 26.9 | 8 | 22.9 | 0.72 |
| Hyperlipidemia | 5 | 8.2 | 3 | 11.5 | 2 | 5.7 | 0.41 |
| Diabetes type 2 | 9 | 14.8 | 2 | 7.7 | 7 | 20.0 | 0.18 |
| Ischemic heart disease | 8 | 13.1 | 2 | 7.7 | 6 | 17.1 | 0.28 |
| Atrial fibrillation | 8 | 13.1 | 3 | 11.5 | 5 | 14.3 | 0.75 |
| Heart failure | 3 | 4.9 | 1 | 3.8 | 2 | 5.7 | 0.74 |
| Asthma | 3 | 4.9 | 0 | 0.0 | 3 | 8.6 | 0.13 |
| COPD | 5 | 8.2 | 3 | 11.5 | 2 | 5.7 | 0.41 |
| Interstitial lung disease | 1 | 1.6 | 0 | 0.0 | 1 | 2.9 | 0.38 |
| Nephropathy | 9 | 14.8 | 5 | 19.2 | 4 | 11.4 | 0.40 |
| Malignancy, organ | 5 | 8.2 | 4 | 15.4 | 1 | 2.9 | 0.07 |
| Malignancy, hematological | 10 | 16.4 | 6 | 23.1 | 4 | 11.4 | 0.07 |
| Autoimmune disease | 6 | 9.8 | 3 | 11.5 | 3 | 8.6 | 0.70 |
| Liver cirrhosis | 1 | 1.6 | 1 | 3.8 | 0 | 0.0 | 0.24 |
| Chronic infection<sup>d</sup> | 2 | 3.3 | 1 | 3.8 | 1 | 2.9 | 0.35 |
| Major surgery within a year | 6 | 9.8 | 2 | 7.7 | 4 | 11.4 | 0.63 |
| Other | 6 | 9.8 | 1 | 3.8 | 5 | 14.3 | 0.18 |
| Neurological comorbidities, any, N (%)<sup>c</sup> | 11 | 18.0 | 5 | 19.2 | 6 | 17.1 | 0.83 |
| Stroke | 3 | 4.9 | 1 | 3.8 | 2 | 5.7 | 0.67 |
| Epilepsy | 1 | 1.6 | 0 | 0.0 | 1 | 2.9 | 0.38 |
| Dementia or MCI | 1 | 1.6 | 1 | 3.8 | 0 | 0.0 | 0.24 |
| Primary headache | 2 | 3.3 | 0 | 0.0 | 2 | 5.7 | 0.46 |
| Neuromuscular | 1 | 1.6 | 1 | 3.8 | 0 | 0.0 | 0.24 |
| Other | 4 | 6.6 | 2 | 7.7 | 2 | 5.7 | 0.76 |
| Psychiatric comorbidities, any, N (%)<sup>c</sup> | 4 | 6.6 | 1 | 3.8 | 3 | 8.6 | 0.46 |
| Depression | 4 | 6.6 | 1 | 3.8 | 3 | 8.6 | 0.46 |
| PTSD | 1 | 1.6 | 0 | 0.0 | 1 | 2.9 | 0.38 |
| Immunosuppression, N (%) | 14 | 23.0 | 9 | 34.6 | 5 | 14.3 | 0.15 |
| **Neurological deficits before admission** |  |  |  |  |  |  | 0.09 |
| Cognitive | 2 | 3.3 | 2 | 7.7 | 0 | 0.0 | - |
| Hemiparesis | 1 | 1.6 | 0 | 0.0 | 1 | 2.9 | - |
| Paraparesis | 1 | 1.6 | 1 | 3.8 | 0 | 0.0 | - |
| Generalized weakness | 2 | 3.3 | 2 | 7.7 | 0 | 0.0 | - |
| Sensory | 1 | 1.6 | 1 | 3.8 | 0 | 0.0 | - |
| **Admission characteristics** |  |  |  |  |  |  |  |
| Nosocomial infection, N (%) | 5 | 8.2 | 0 |  | 5 | 14.3 | 0.04 |
| Symptom debut to admission, days, mean (SD) | 7.2 | 8.5 | 7.5 | 7.8 | 7 | 9.2 | 0.82 |
| Admission duration, days, mean (SD) | 29.7 | 21.6 | 17.5 | 13.9 | 38.9 | 22.0 | <0.001 |
| Dialysis | 17 | 27.9 | 0 | 0 | 17 | 48.6 | <0.001 |
| ICU admission, N (%) | 35 | 57.4 | - | - | - | - | - |
| Mechanical ventilation<sup>e</sup> | 34 | 97.1 | - | - | - | - | - |
| ECMO<sup>e</sup> | 6 | 17.1 | - | - | - | - | - |
| Days in ICU, mean (SD) | 19 | 12.2 | - | - | - | - | - |

Basic demographics, past medical history and admission characteristics of COVID-19 positive patients dichotomized in ICU and non-ICU patients.
<sup>a</sup> For categorical values we tested with Fisher's exact test; for normal distributed values we used a student's t-test and for non-normal distributed values we calculated median and tested with non-parametric test for p-value.
<sup>b</sup> If not stated then the percentage is of the total population.
<sup>c</sup> One patient may have several comorbidities.
<sup>d</sup> Two patients with chronic infection refers to Hepatitis B and long-term Clostridium Difficile infection.
<sup>e</sup> Percentage of total ICU patients.
ICU, intensive care unit; SD, standard deviation; IQR, interquartile range; COPD, chronic obstructive pulmonary disease; mRS, modified Rankin scale; MCI, mild cognitive impairment; PTSD, post-traumatic stress disorder; ECMO, extracorporeal membrane oxygenation.

**Table 2.**
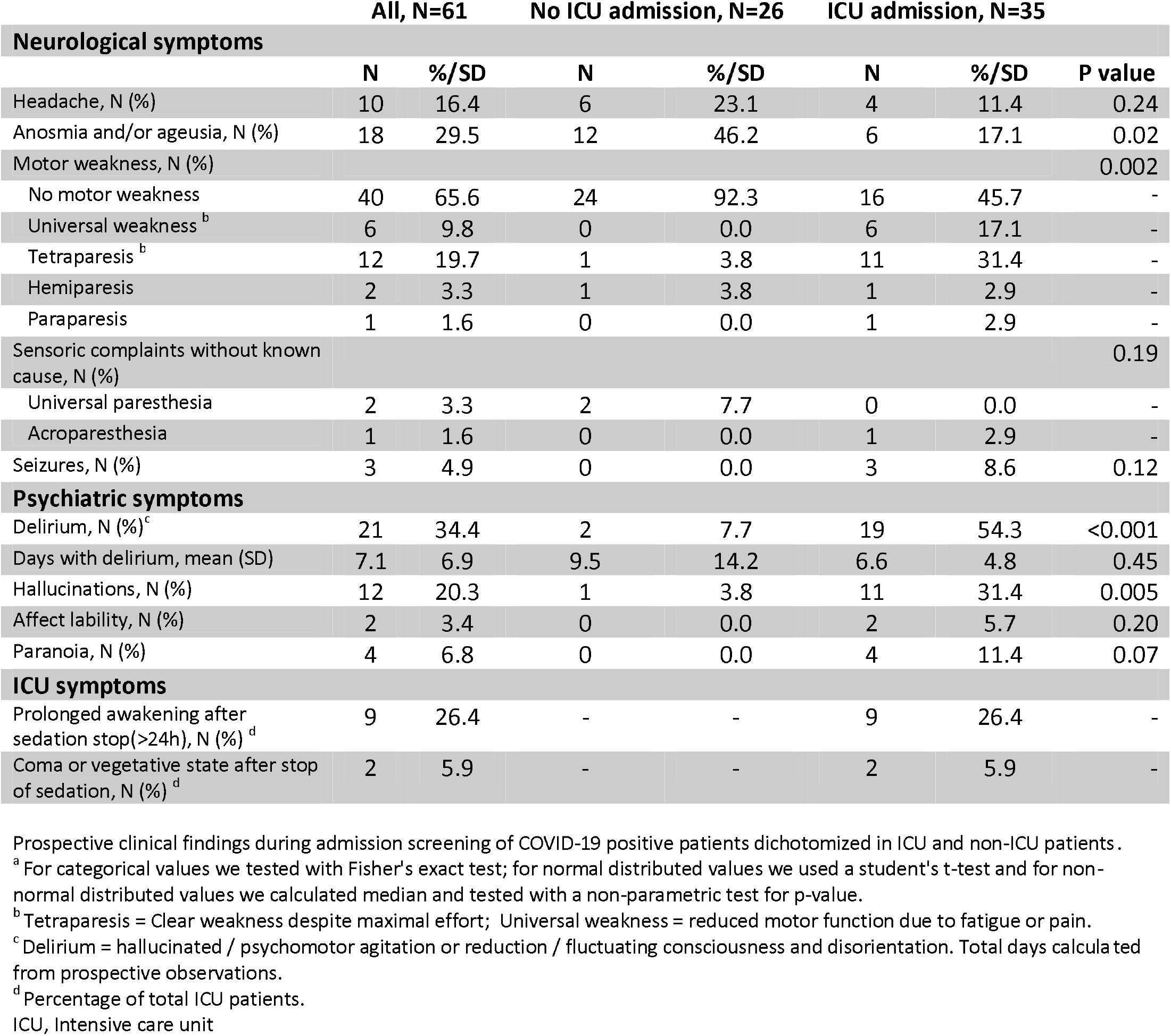
Symptoms and signs during admission ^a^.

**Table 3.** Discharge evaluation ^a^.

|  | All, N=61 |  | No ICU admission, N=26 |  | ICU admission, N=35 |  | P value |
| --- | --- | --- | --- | --- | --- | --- | --- |
| In-hospital death, N (%) | 10 | 16.4 | 1 | 3.8 | 9 | 25.7 | 0.017 |
| mRS at discharge, median (IQR) | 3 | 2-5 | 2 | 1-3 | 4 | 4-6 | <0.001 |
| MOCA test performed, N (%) | 32 | 52.5 | 18 | 69.2 | 14 | 40.0 | 0.03 |
| MOCA score, median (IQR) | 22 | 18.5-28 | 25 | 22-29 | 19.5 | 16-24 | 0.02 |
| Full neurological examination performed, N (%)<sup>b</sup> | 53 | 86.9 | 26 | 100 | 27 | 77.1 |  |
| **Neurological deficits at discharge, N (%) <sup>c</sup>** |  |  |  |  |  |  |  |
| **Cognitive** |  |  |  |  |  |  |  |
| Amnesic | 18 | 34.0 | 5 | 19.2 | 13 | 48.1 | 0.01 |
| Aphasia | 2 | 3.8 | 0 | 0.0 | 2 | 7.4 | 0.12 |
| Apraxia | 4 | 7.5 | 2 | 7.7 | 2 | 7.4 | 0.9 |
| Dysexecutive | 20 | 37.7 | 5 | 19.2 | 15 | 55.6 | 0.001 |
| Neglect | 0 | 0.0 | 0 | 0.0 | 0 | 0.0 | - |
| **Motor and gait** |  |  |  |  |  |  |  |
| Hemiparesis | 2 | 3.8 | 1 | 3.8 | 1 | 3.7 | - |
| Paraparesis | 2 | 3.8 | 1 | 3.8 | 1 | 3.7 | - |
| Tetraparesis | 11 | 20.8 | 0 | 0.0 | 11 | 40.7 | <0.001 |
| Tetraparesis with hyporeflexia and atrophy | 8 | 15.1 | 0 | 0.0 | 8 | 29.6 | 0.002 |
| Generalized weakness | 6 | 11.3 | 1 | 3.8 | 5 | 18.5 | - |
| Gait impairment, assisted with walking device | 11 | 20.8 | 5 | 19.2 | 6 | 22.2 | - |
| Gait impairment, bedridden/wheelchair | 12 | 22.6 | 1 | 3.8 | 11 | 40.7 | 0.001 |
| Ataxia and/or dysmetric coordination | 7 | 13.2 | 0 | 0.0 | 7 | 25.9 | 0.02 |
| Dysarthria and/or dysphagia | 3 | 5.7 | 0 | 0.0 | 3 | 11.1 | 0.16 |

Discharge evaluation of COVID-19 positive patients dichotomized in ICU and non-ICU patients.
<sup>a</sup> For categorical values we tested with Fisher's exact test; for normal distributed values we used a student's t-test and for non-normal distributed values we calculated median and tested with non-parametric test for p-value.
<sup>b</sup> Patients who died during heavy sedation or were transferred to other ICU at different hospitals were not examined at discharge.
<sup>c</sup> One patient may have several deficits.
ICU, Intensive care unit; mRS; Modified Rankin Scale; IQR, Inter quartile range; MOCA, Montreal cognitive assessment.

**Table 4.** Follow-up at 3 months ^a^.

|  | All, N=45<sup>b</sup> |  | No ICU admission, N=21 |  | ICU admission, N=24 |  | P value |
| --- | --- | --- | --- | --- | --- | --- | --- |
| Death after discharge, N (%) | 2 | 4.4 | 2 | 9.5 | 0 | 0 | 0.12 |
| Hospitalization after discharge total, N (%) | 17 | 37.8 | 9 | 42.9 | 8 | 33.3 | 0.04 |
| Neurological department | 4 | 23.5 | 0 | 0 | 4 | 50 | - |
| Psychiatric department | 0 | 0 | 0 | 0 | 0 | 0 | - |
| Other | 13 | 76.5 | 9 | 100 | 4 | 50 | - |
| 3-month mRS, median (IQR) | 1 | 1-2 | 1 | 1-2 | 1 | 0.5-2.5 | 1.00 |

Follow-up data 3 months after discharge of COVID-19 positive patients dichotomized in ICU and non-ICU patients.
<sup>a</sup> For categorical values we tested with Fisher's exact test; for normal distributed values we used a student's t-test and for non-normal distributed values we calculated median and tested with non-parametric test for p-value.
<sup>b</sup> Follow-up data on 45 patients. Ten patients died during admission; 6 patients still not passed 3 months after discharge.
mRS, Modified Rankin Scale; IQR, Inter quartile range; ICU, Intensive care unit.

**Table 5.** Laboratory findings ^a^.

|  | All, N=61 |  | No ICU admission, N=26 |  | ICU admission, N=35 |  | P value |
| --- | --- | --- | --- | --- | --- | --- | --- |
| **CT Brain performed, N (%)** | 16 | 26.2 | 4 | 15.4 | 12 | 34.3 | 0.08 |
| **CT Brain findings, N (%)** |  |  |  |  |  |  | 0.09 |
| Normal or slight atrophy | 7 | 42.9 | 0 | 0.0 | 7 | 54.5 | - |
| Old stroke | 3 | 21.4 | 1 | 25.0 | 2 | 18.2 | - |
| Leukoaraiosis | 1 | 7.1 | 1 | 25.0 | 0 | 0.0 | - |
| New ischemic stroke | 1 | 7.1 | 1 | 25.0 | 0 | 0.0 | - |
| Hemorrhagic stroke | 1 | 7.1 | 0 | 0.0 | 1<sup>b</sup> | 9.1 | - |
| Superficial siderosis / Subarachnoid Bleed | 1 | 7.1 | 0 | 0.0 | 1 | 9.1 | - |
| Other | 2 | 14.2 | 1 | 25.0 | 1 | 9.1 | - |
| **MRI performed, N (%)** | 7 | 11.4 | 4 | 15.4 | 3 | 8.6 | 0.73 |
| **MRI findings, N (%)** |  |  |  |  |  |  | 0.41 |
| Normal | 0 | 0 | 0 | 0 | 0 | 0 | - |
| New ischemic stroke | 1 | 16.7 | 1<sup>c</sup> | 25.0 | 0 | 0 | - |
| Inflammatory lesions | 1 | 16.7 | 1<sup>d</sup> | 25.0 | 0 | 0 | - |
| Hydrocephalus | 1 | 16.7 | 0 | 0 | 1 | 33.3 | - |
| Atrophy | 2 | 33.3 | 1 | 25.0 | 1 | 33.3 | - |
| Other | 2 | 33.3 | 1 | 25.0 | 1 | 33.3 | - |
| **EEG performed, N (%)** | 12 | 19.7 | 2 | 7.7 | 10 | 28.6 | 0.04 |
| **EEG findings, N (%)** |  |  |  |  |  |  | 0.89 |
| Normal | 0 | 0 | 0 | 0 | 0 | 0 | - |
| FIRDA | 5 | 41.7 | 1 | 50 | 4 | 40 | - |
| Sharp waves | 1 | 8.3 | 0 | 0 | 1 | 10 | - |
| Encephalopathic curve | 6 | 50 | 1 | 50 | 5 | 50 | - |
| Epileptic activity | 0 | 0 | 0 | 0 | 0 | 0 | - |
| **LP performed, N (%)** | 8 | 13.1 | 3 | 11.5 | 5 | 14.3 | 0.72 |
| **CSF findings, N (%)** |  |  |  |  |  |  | 0.19 |
| Normal cell count | 5 | 62.5 | 1 | 33.3 | 4 | 80 | - |
| CSF pleocytosis (>5 cells/microl) | 3 | 37.5 | 2<sup>c,d</sup> | 66.6 | 1<sup>e</sup> | 20 | - |
| Cell levels (E6/L), median (IQR) | 3 | 2.5-69 | 125 | 1-271 | 3 | 3-3 | 0.45 |
| Protein levels (g/L), median (IQR) | 0.53 | 0.37-0.86 | 0.59 | 0.43-1.13 | 0.49 | 0.3-0.56 | 0.46 |
| SARS-CoV-2 CSF PCR performed (positive) | 5 | 0 | 2 | 0 | 3 | 0 | - |
| **ENG / EMG performed, N (%)** | 1 | 1.6 | 0 | 0 | 1 | 2.9 | 0.38 |
| **ENG / EMG Findings, N (%)** |  |  |  |  |  |  | - |
| Critical illness myopathy | 1 | 100 | 0 | 0 | 1 | 100 | - |

Paraclinical findings of COVID-19 positive patients dichotomized ICU and non-ICU patients.
<sup>a</sup> For categorical values we tested with Fisher's exact test for p-value.
<sup>b</sup> Patient initially admitted with cerebellar ICH. COVID-19 infection during hospitalization.
<sup>c</sup> One patient with multiple strokes, CSF pleocytosis and severe encephalopathy (see figure 2 for case and images).
<sup>d</sup> Patient with transverse myelitis seen on spinal MRI and with CSF pleocytosis (see figure 2 for case and images).
<sup>e</sup> Severe headache and signs of CNS infection at admission. Positive VZV antibodies in CSF and pleocytosis with 13 cells.
ICU, Intensive care unit; CT, computed tomography; MRI, magnetic resonance imaging; EEG, electroencephalogram; LP, lumbar puncture; ENG, electroneurography; EMG, electromyography.

**Table 6.** Diagnoses based on complications during admission and after 3 months ^a^.

|  | All, N=61 |  | During admission, N=61 |  | After discharge, N=45 |  |
| --- | --- | --- | --- | --- | --- | --- |
| **Cerebrovascular, N (%)** | **N** | **%** | **N** | **%** | **N** | **%** |
| Stroke, any type | 4 | 6.6 | 2 | 3.3 | 2 | 4.4 |
| Ischemic | 4 | 100 | 2 | 100 | 2 | 100 |
| Hemorrhagic | 0 | 0 | 0 | 0 | 0 | 0 |
| **Central nervous system, N (%)** |  |  |  |  |  |  |
| Encephalopathy, any | 19 | 31.1 | 19 | 31.1 | 1<sup>e</sup> | 2.2 |
| Mild encephalopathy | 6 | 31.6 | 6 | 31.6 | 0 | - |
| Severe encephalopathy | 13 | 68.4 | 13 | 68.4 | 1<sup>e</sup> | - |
| Severe encephalopathy with akinetic mutism | 8 | 42.1 | 8 | 42.1 | 0 | - |
| Encephalitis | 2<sup>b,c</sup> | - | 2 | - | 0 | - |
| Myelitis | 1<sup>d</sup> | - | 0 | - | 1 | - |
| Other | 1<sup>f</sup> |  | 0 |  | 1<sup>f</sup> |  |
| **Peripheral nervous system, N (%)** |  |  |  |  |  |  |
| Neuropathy, any | 12 | 19.7 | 8 | 13.1 | 4 | 8.9 |
| Peripheral facial palsy | 2 | 16.7 | 0 | 0 | 2 | 50 |
| Meralgia paresthetica | 2 | 16.7 | 0 | 0 | 2 | 50 |
| CIPM, definite | 1 | 8.3 | 1 | 8.3 | 0 | 0 |
| CIPM, possible | 7 | 58.3 | 7 | 58.3 | 0 | 0 |
| Guillain Barre syndrome | 0 | 0.0 | 0 | 0 | 0 | 0 |
| Acute myopathy, any | 1 | 8.3 | 1 | 8.3 | 0 | 0 |
| Other | 1<sup>g</sup> | 8.3 | 0 | 0 | 1<sup>g</sup> | 2.2 |

Cerebrovascular, Central and peripheral nervous system complications during admission and after discharge of COVID-19 positive patients.
<sup>a</sup> One patient may have more than one complication
<sup>b</sup> Acute Hemorrhagic Necrotizing Encephalopathy (see figure 2 for image and case)
<sup>c</sup> One patient with severe apathy and amnesia without focal deficits, multiple ischemic lesions on MRI and CSF pleocytosis (see figure 2 for image and case)
<sup>d</sup> Patient with transverse myelitis seen on spinal MRI and with CSF pleocytosis.
<sup>e</sup> One patient developed severe agitation, fits of screaming and automatisms with rocking back and forward. Psychiatric evaluation at nursing home without explanation. Treated with pregabalin that calmed the symptoms. The patient did also develop severe encephalopathy during admission. Lumbar puncture with normal cell and protein levels.
<sup>f</sup> Admitted from a training facility with recurrent generalized seizures. CT of the brain and CSF investigation with normal findings. Severe hypomagnesemia due to possible refeeding syndrome.
<sup>g</sup> Weeks after discharge, one patient developed right sided drop foot, facial palsy, mild dysphagia and hypoglossal palsy. MRI normal, ENG with bilateral mild peroneal affection without specification. Symptoms resolved spontaneously CIPM; Critical illness poly-neuropathy or myopathy.

**Figure 1.**
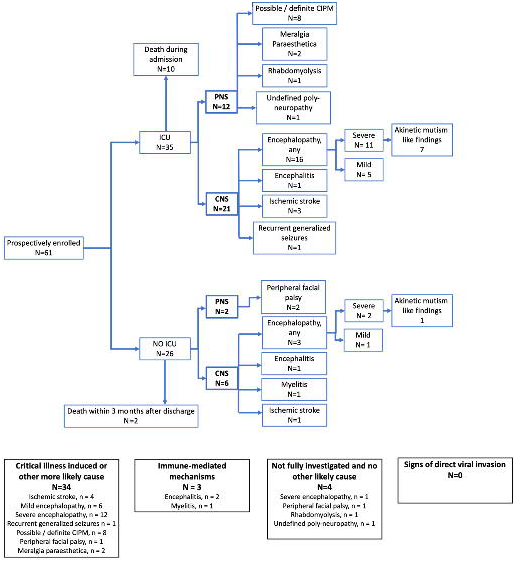
Flow-chart of patient inclusion and observed neurological complications. Prevalence of prespecified complications observed in 61 prospectively enrolled COVID-19 patients, hospitalized in a tertiary center from April 1 to September 25, 2020. Patients were dichotomized in ICU and non-ICU patients. The observation period was from admission to 3-month after discharge or death. N = number of patients with the specified complication, one patient may have several complications. CNS; central nervous system. PNS; peripheral nervous system. CIPM; critical illness polyneuromyopathy. MP; meralgia paresthetica. ICU; intensive care unit.

**Figure 2.**
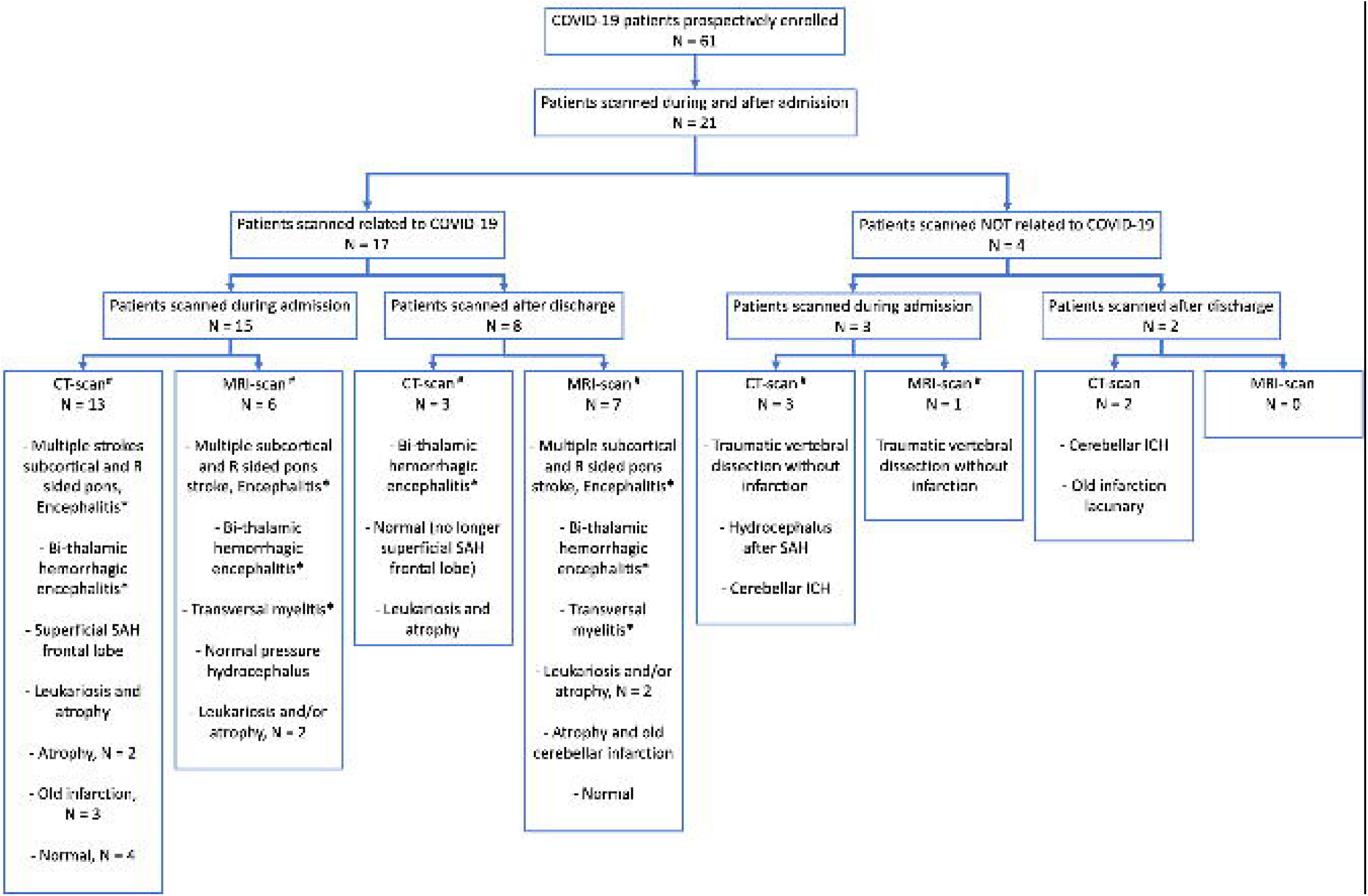
Flow-chart of brain imaging performed during and after admission. Brain imaging overview of 61 prospectively enrolled COVID-19 patients from admission to 3 month after discharge, dichotomized into COVID-19-related scans referring to neuroimaging related to a COVID-19 suspected complication or a complication related to a COVID-19 admission and neuroimaging performed for other reasons (e.g. traumatic injury) or for disease that occurred before COVID-19 (e.g. cerebral hemorrhage with onset before COVID-19). All images were re-evaluated by two board-certified neuroradiologists. # One patient may be scanned multiple times with both CT and MRI and represented in both categorized groups of scanned during and after admission. * See Figure 2 for images and case history. R; right. ICH; intracerebral hemorrhage. SAH; subarachnoid hemorrhage. CT; computed tomography. MRI; magnetic resonance imaging.

### 3.1 Clinical screening during admission

#### 3.1.1 Neurological symptoms

The most common neurological finding was motor weakness seen in 21 patients (34.4%) which was significantly more prevalent in ICU-patients (54%) compared to non-ICU patients (7.7%), p=0.002. The second most common finding was anosmia and/or ageusia in 18 patients (29.5%), significantly more prevalent in non-ICU patients (46.2%) compared to ICU-patients (17.1%), p=0.02. Ten patients (16.4%) complained of headache and 3 (4.9%) of sensory symptoms, which were not attributable to a specific cause. Seizures were observed in 3 patients (4.9%), including epileptic seizures in 2 of these patients (generalized tonic-clonic seizures), and non-epileptic seizures in 1 patient with dystonic painful contractions/seizures in the right hand lasting 10-20 seconds every day for 6 days. Unfortunately, no brain imaging or EEG was performed in the latter patient because he deteriorated in respiratory function and died in the ICU.

Of the 35 ICU-admitted patients, 9 (26.4%) had prolonged awakening from sedation and clinical unresponsiveness in >24h, resulting in EEG investigations in 7 patients. Two patients (5.9%) were in prolonged coma off sedation, resulting in death due to withdrawal of life-supporting therapy following discussions with the families.

#### 3.1.2 Neuropsychiatric symptoms

A total of 21 (34.4%) patients had delirium during admission lasting 7.1 days (mean, standard deviation (SD) 6.9), which was significantly more frequent in ICU-patients (54.3%) compared to non-ICU patients (7.7%), p<0.001. The most common finding in delirious patients was hallucinations (12 patients, 20.3%). **Table 2** shows a full outline of CNS and PNS symptoms during admission.

#### 3.1.3 Laboratory investigations

During admission, CT was performed in 16 patients (26.2%), MRI in 7 patients (11.4%), EEG in 12 patients (19.7%), and CSF was investigated in 8 patients (13.1%). ICU-admitted patients were more frequently investigated with EEG than non-ICU patients (28.6% vs. 7.7%, p=0.04), and the most common findings were compatible with an encephalopathic curve, i.e. background slowing including increased theta and delta activity, seen in 50% of all the investigated patients, and frontal intermittent rhythmic delta activity (FIRDA) seen in 40% of the investigated patients. Of 8 patients who had CSF investigations during the study period, 3 patients had pleocytosis (cell count >5 cells/microl): In one patient with severe headache and respiratory failure, a lumbar puncture showed 13 cells/microl with microbiological investigations revealing intrathecal varicella zoster antibodies, but CSF SARS-CoV-2 PCR was not performed. The other 2 patients with CSF pleocytosis had myelitis and encephalitis (see 3.4 for details). Five of the 8 CSF samples were tested for SARS-CoV-2 RNA, and all were negative. For full outline of paraclinical findings see **Table 5** and **Figure 2**.

### 3.2 Clinical screening at discharge

Of the 61 patients, 53 (86.9%) were examined at discharge with a full neurological exam, including cognitive screening for amnesia, dysexecutive function, apraxia, aphasia and neglect; and 32 patients (52.5%) were examined with a MOCA. As already stated, 10 patients died during their hospital stay (16.4%).

#### 3.2.1 Neurological symptoms

We observed tetraparesis with hyporeflexia and muscle atrophy in 8/53 (15%) and gait impairment categorized as bedridden/wheelchair in 12/53 (22.6%) patients, which both were significantly more frequent in ICU patients, p=0.002 and p=0.001, respectively. Median MOCA score was 22 in the total population, but lower in patients admitted to the ICU (median 19.5) than in non-ICU patients (median 25), p-value = 0.02. Median mRS at discharge was 3 (IQR 2-5). See **Table 3** for full data on discharge evaluation.

#### 3.2.2 Neuropsychiatric symptoms

Amnesia (n=18/53, 34.0%) and dysexecutive function (20/53, 37.7%) were the most common cognitive deficits and more prevalent in ICU patients compared to non-ICU patients, p=0.01 and p<0.001, respectively.

### 3.3 Follow-up after 3-month

Three-month follow-up data was available for 45 patients (**Table 4**). The median 3-month mRS was 1 (IQR 1-2), which was not significantly different between ICU-admitted (mRS=1, IQR: 0.5-2.5) and the remaining patients (mRS=1, IQR: 1-2). Seventeen patients (37.8%) were re-hospitalized after discharge, and 4 of these (23.5%) were re-hospitalized at a neurological department: One patient had a new-onset ischemic stroke; 1 had recurrent generalized tonic-clonic seizures due to hypomagnesemia secondary to a refeeding syndrome; 1 had memory decline and gait difficulties with brain MRI showing normal pressure hydrocephalus; and finally, 1 patient had new-onset mild dysphagia, tongue palsy and a right sided drop-foot, compatible with hypoglossal and peroneal nerve injuries, investigated with a brain MRI with normal findings and ENG showing bilateral mild peroneal nerve affection; symptoms resolved spontaneously after 3 weeks. Follow-up data was missing for 16 patients (10 deaths, 6 patients with less than 3-month follow-up).

### 3.4 CNS and PNS complications during admission and at 3-month follow-up

Based on the definition of complications as outlined in section 2.5 (see Methods) and prospective observations during admission, discharge and at 3-month follow-up, we observed and diagnosed the complications outlined below and in **Table 6**.

#### 3.4.1 Cerebrovascular

Four patients had new-onset ischemic stroke, 2 patients during their COVID-19 admission and 2 patients between discharge and 3-month follow-up. Mean age of the patients was 75 years (SD 3.1), time from COVID-19 symptom debut to stroke onset was 40.5 days (median), and all patients had one or several cardiovascular risk factors.

#### 3.4.2 CNS

A total of 19 patients (31.1%) had signs of encephalopathy at some stage during their admission, and 13 (68.4%) of those were categorized as having severe encephalopathy (GCS 12 or less). Patients with severe encephalopathy had significantly longer hospital stays (median 21 vs. 44 days without encephalopathy, p <0.001), required more often treatment with haloperidol (61.5% vs. 16.7% with less severe encephalopathy, p=0.03) and were more often admitted to the ICU (80% vs. 50% of all patients, p=0.04). Among the 13 patients with severe encephalopathy, we encountered 8 patients who were apathetic and severely hypokinetic/akinetic, with almost no voluntary movements and without any verbal output, yet they were able to follow the examiner with the eyes and indicate yes and no to questions by nodding or shaking their heads. On examination, all these patients had a palmomental reflex. This state of akinetic mutism lasted for a median of 4 days (IQR 2.5-6.5). Seven of the 8 patients with akinetic mutism had been admitted to the ICU, 5 required dialysis for renal failure, 4 were treated with haloperidol for delirium, 3 had FIRDA on EEG, 1 had multiregional sharp waves on EEG, and 1 patient had a non-traumatic cortical subarachnoid bleed in the right frontal lobe.

Two patients were diagnosed as having encephalitis. One patient, previously healthy, was admitted with generalized epileptic seizures 10 days after onset of fever, dry cough and myalgia and a strongly positive IgG antibody response to SARS-CoV-2 (**Figure 3a**). Brain MRI showed bilateral basal ganglia ischemic/inflammatory lesions with hemorrhagic transformation, and CSF showed normal cell and protein count; CSF SARS-CoV-2 PCR was negative. A brain biopsy was obtained to rule out a brain abscess, but investigations for a microbiological, fungal or neoplastic etiology were negative. Biopsy showed multiple fibrinous thrombosis in the microvasculature in keeping with a histological diagnosis of hemorrhagic necrotizing encephalitis. The patient was diagnosed as having COVID-19 associated acute necrotizing encephalitis and referred for neurorehabilitation.

**Figure 3.**
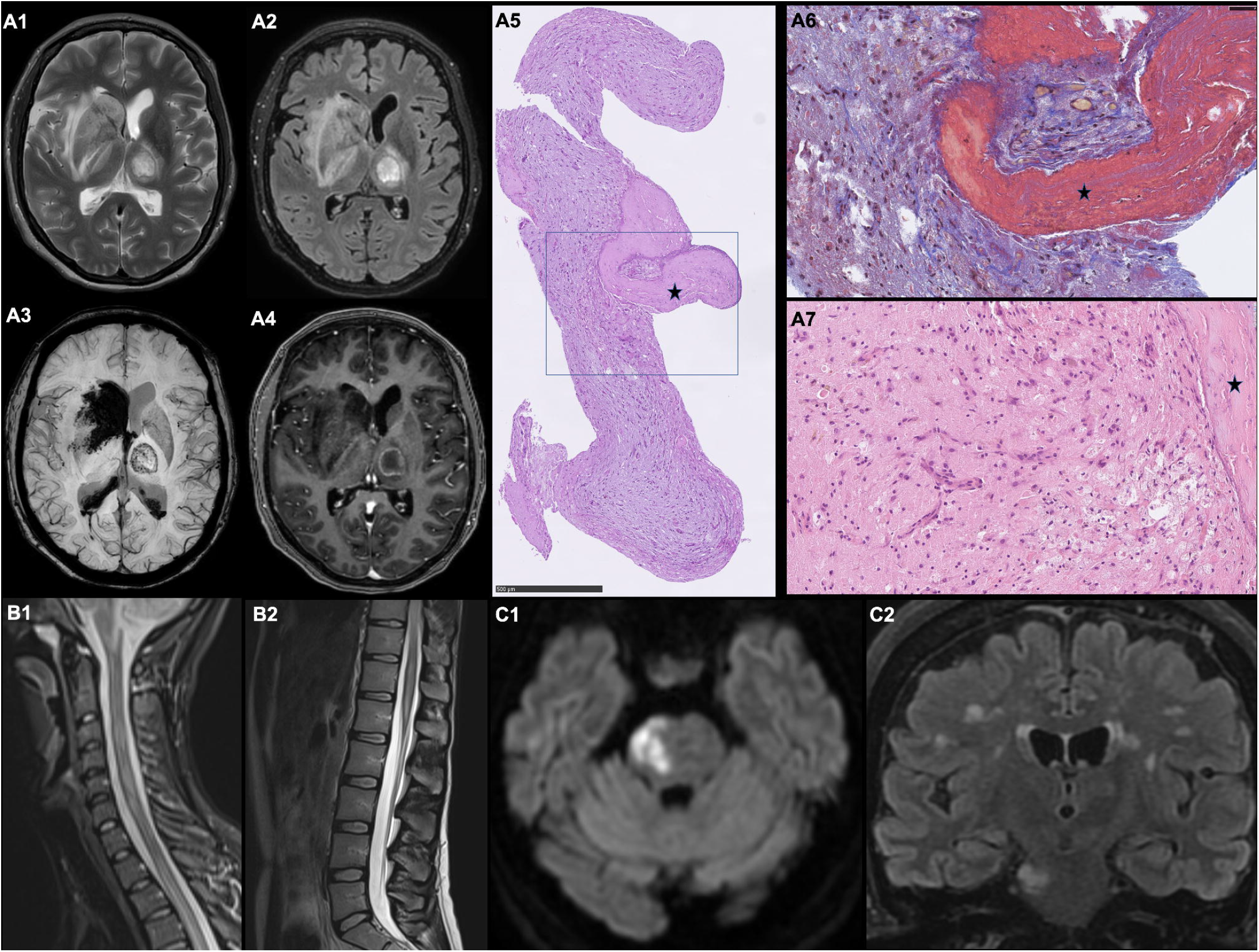
Brain imaging and histopathology from probable COVID-19 encephalitis and myelitis. **A**. 50+-year-old patient was admitted with convulsive seizures and reduced consciousness 19 days after symptom onset with dry cough, fever, a sore throat, headache and myalgias. Three weeks prior to presentation her entire family had experienced flu-like symptoms and a family member was tested positive for SARS-CoV-2. Previous medical history included hypertension, heterozygous Factor V Leiden mutation and chronic tension type headache. Medication before admission included amlodipine and gabapentin. On the morning of admission, the patient had generalized convulsive seizures. On examination, right-sided eye deviation and aphasia were noted. Twenty-four hours after admission, owing to a sudden decrease in consciousness and progressive hypoxemic respiratory failure, the patient was intubated and transferred to the ICU. Chest X-ray revealed a complete white-out of the left lung. Microscopic examination of the bronchoalveolar lavage fluid revealed no microorganisms, and the BAL fluid was negative with broad infectious workup, including culture, tests for SARS-CoV-2, influenza virus type A and B, adenovirus, metapneumovirus, parainfluenza virus, atypical pneumonia, mycobacteria, pneumocystis, fungi and CMV, except for a positive aspergillus galactomannan antigen. Cytologic examination of the lung biopsy revealed type II pneumocyte hyperplasia and capillary inflammation with macrophages and neutrophils. No hyaline membranes, granulomas, necrosis, fibrosis or vasculitis were observed. There was no microscopic evidence of malignancy or fungi, including immunohistochemical (IHC) staining for fungi. In the alveoli, there were small precipitations of fibrin. The findings were concluded to be unspecific, resembling findings observed in viral infection. Assays for antinuclear antibodies, anti-double-stranded DNA and rheumatoid factor were negative. Lupus anticoagulant and anticardiolipin antibodies were negative. Electroencephalography revealed generalized slowing, with no epileptiform activity and brain MRI demonstrated on axial T2W (*A1*) and FLAIR (*A2*) bilateral hyperintense lesions in the right basal ganglia and left thalamus. Axial SWI images (*A3*) demonstrated hemorrhage, seen as hypointense signal intensity in the right basal ganglia lesion and ventricle system. Microbleeds were seen in the left thalamic lesion (*A3*) and ring enhancement on the post contrast T1W images (*A4*). A lumbar puncture was performed. Gram’s staining of the cerebrospinal fluid (CSF) showed no cells or organisms. The white-cell count was 0 per microliter (reference range, <5), the red-cell count was 0 per microliter (reference range, <300), the glucose level was 4.4 mmol per liter (2.2 to 3.9 mmol per liter) and the protein level was 0.3 g per liter (reference value, 0.15 - 0.50). Cytologic examination of the CSF revealed no malignant cells, and there was no evidence of B-of T-cell lymphoproliferative disorder on flow cytometry. Polymerase-chain-reaction assays of the CSF for herpes simplex virus and VZV were negative, as were mycobacterial smear and culture of the CSF. Culture of the CSF showed no growth. Testing of the CSF for autoantibodies, Lyme disease, syphilis, human herpes virus and SARS-CoV-2 were negative. Whole-body positron-emission tomography and computed tomography (PET-CT), performed after the administration of intravenous 18F-fluorodeoxyglucose (FDG) tracer, revealed no suspicion of extracranial malignant disease. The differential diagnosis consisted of cerebral abscess or malignant tumor, and the patient was transferred to the neuro-ICU for further evaluation and treatment. To obtain a neuropathological diagnosis, stereotactic biopsy of the cerebral lesions was performed (*A5-7*). Biopsy specimens from both right and left cerebral hemispheres showed thrombotic occlusion of small vessels by fibrin thrombi. The brain parenchyma showed reactive changes with astrogliosis and microgliosis, axonal swellings, proliferating capillaries and iron deposits (Perl’s stain) as sign of microbleeds. There were a sparse diffuse lymphocytic infiltrate consisting of CD3 positive T-lymphocytes. CD20 staining showed no B-lymphocytic infiltrates. There were no granulomas. There was no sign of demyelinating disease when comparing neurofilament immunostaining with myelin stains (LUXPAS and immunostaining for myelin basic protein). Immune staining’s for Toxoplasma, Herpes 1, Cytomegalovirus and JC virus were all negative. There were no fungi or bacteria in special staining’s with PAS, Gomori and Gram. Biopsy from the left cerebral hemisphere (*A5*) with thrombi and reactive changes (PAS staining). Fibrin thrombus in a small vessel (Masson trichrome staining) (*A6*). Higher magnification showing proliferating capillaries, hemosiderin in macrophages and gliosis (hematoxylin and eosin stain), (*A7*). Asterix: thrombus. The team of neuropathologists concluded the findings in the brain biopsy tissue to be consistent with small necrotic lesions/infarctions, likely caused by fibrin thrombi in the vessels. The clinical suspicion of COVID-19 was confirmed by a positive serum test for SARS-CoV-2 showing high titers of neutralizing IgG-antibodies obtained on the 27th day of illness. A diagnosis of COVID-19-associated acute necrotizing encephalitis was made based on clinical signs, MRI, brain pathology findings and high titers of SARS-CoV-2 IgG and exclusion of other causes following extensive diagnostic work-up. **B**. A 20+-year-old patient with a previous medical history of hypothyroidism developed a dry cough and myalgia in the beginning of March and was tested SARS-CoV-2 positive 3 days later. COVID-19 symptoms lasted 6 days. Eight days after onset of COVID-19 symptoms, the patient developed lumbar back pain and progressive numbness in both legs with a mid-thoracic sensory level. The patient further developed paresthesia in the right hand, difficulties climbing stairs, and urinary retention with a sense of constant urge. Neurological examination revealed a mild symmetric tetraparesis, hypoesthesia in both legs with a Th5 dermatome sensory level, impaired proprioception in the legs, absent abdominal reflexes and a slightly broad-based gait. Cognitive evaluation and the rest of the exam were unremarkable. Spinal MRI demonstrated on sagittal T1 STIR (*B1*) and sagittal T2 (*B2*) hyperintense lesions from C1 to C7 and from Th1 to the conus medullaris, consistent with transverse myelitis. Lumbar puncture showed a lymphocytic pleocytosis of 125 cells/microl, protein levels of 0.59 g/L with a negative microbiological cerebrospinal fluid work-up including herpes simplex virus, varicella zoster virus, cytomegalovirus, enterovirus, and SARS-CoV-2. Blood was negative for HIV and syphilis. Screening for aquaporin-4 and myelin oligodendrocyte glycoprotein antibodies was negative. The patient was treated with high-dose prednisone and plasma exchange and made an uneventful recovery. Eight months later, the patient had a mild degree of Lhermitte’s but no other complaints. **C**. A 60+-year-old patient with a previous medical history of type 2 diabetes, hypothyroidism and kidney transplant in 2019 due to adult onset polycystic kidney disease and immunosuppression with ciclosporine, developed muscle ache, dry cough, fever and dizziness resulting in a positive SARS-CoV-2 pharyngeal swab test and admission for 6 days. Eleven days after COVID-19 symptom onset, the patient developed headache, progressive confusion, apathy and speech difficulties and was re-admitted. Neurological examination revealed the patient was apractic, apathetic and amnestic. She answered questions with latency and with a mild, non-fluent aphasic speech. Brain MRI demonstrated on axial DWI (*C1*) a hyperintense lesion in the right side of pons and on coronal FLAIR (*C2*) multiple sub- and juxta-cortical hyperintense lesions. Lumbar puncture showed a lymphocytic pleocytosis of 252 cells (E6/L), protein levels of 1.77 g/L. CSF microscopy was unremarkable, and cultures for bacteria and fungi were negative. CSF screening for E.coli., Hemophilus influenzae, listeria, neisseria, streptococcus, enterovirus, Cryptococcus neoformans, syphilis (both intrathecal and peripheral testing), Lyme’s disease, JC virus, 18S-RNA, flow cytometry, and LGI1, NMDA, GABA-B, and CASPR2 antibody screening for autoimmune encephalitis were all negative. HIV and hepatitis B testing, transesophageal echocardiography, whole body FDG-PET for occult malignancy were also unremarkable. There were a positive PCR and intrathecal antibody titer for EBV, HSV-1, VZV and human herpes virus but the attending infectious disease and microbiology colleagues considered this to be consistent with a reactive phenomenon owing to chronic immunosuppression. The patient was initially treated for a few days with ceftriaxone and acyclovir, but treatment was stopped when no clear evidence of bacterial or herpes virus encephalitis was found. The patient made a gradual and spontaneous recovery, but cognitive impairment of memory and executive function remained after discharge.

The second patient with encephalitis had a medical history of kidney transplantation and immunosuppression and was admitted due to myalgia and a dry cough with a positive SARS-CoV-2 PCR on pharyngeal swab and, 11 days later, developed severe apathy, amnesia and initially mild aphasia but no other focal deficits (**Figure 3b**). Brain MRI showed multiple ischemic lesions bilaterally in the basal ganglia, thalami, juxtacortical white matter and in the right side of the pons. CSF analysis revealed lymphocytic pleocytosis (250 cells/microl), and microbiological screening showed positive EBV, HSV-1 and VZV DNA copies (which the attending microbiologist concluded was reactive and associated with chronic immunosuppressive therapy). SARS-CoV-2 PCR on CSF material was negative, but SARS-CoV-2 antibodies were positive in the CSF. The patient was discharged to neurorehabilitation and improved slowly with a decline in CSF cell count on follow-up without specific treatment.

One patient developed lumbar pain, thoracic numbness with a sensory level at T5, urinary retention and mild paraparesis 13 days after COVID-19 symptom onset with myalgia and fever, and 6 days after remission of COVID-19 symptoms (**Figure 3c**). Spinal MRI showed transverse myelitis, CSF analysis revealed 125 lymphocytic cells/microl, and microbiological screening including SARS-CoV-2 PCR was negative. The patient made an uneventful recovery following steroid treatment and plasma exchange.

We used a logistic regression model with the covariates age, sex, number of admission days number of days in the ICU, dialysis, delirium, and treatment with haloperidol. We found that duration of ICU admission was an independent predictor for severe encephalopathy with an increase of OR=1.23 (95% CI=1.03-1.47) for each day in the ICU, p=0.02.

#### 3.4.3 PNS

During admission, we observed 9 (13.1%) patients with PNS complications. Eight patients were tetraparetic at discharge, had muscle atrophy and hyporeflexia, and all had been admitted to the ICU. One of these 8 patients was investigated with ENG/EMG and diagnosed with definite CIPM, the remainder were categorized as having possible CIPM. All 8 patients with definite or possible CIPM, when compared to patients without neuromuscular complications, had significantly longer admissions (median 49.5 vs. 23 days, p<0.001), were more frequently admitted to the ICU (100% vs. 40%, p=0.002), more often treated with renal replacement therapy (62% vs. 14%, p=0.008), and were more often delirious (75% vs. 30%, p=0.02).

Further, one patient was admitted with rhabdomyolysis and kidney failure with a peak creatine kinase of 74.000 Units/L. Following symptomatic treatment including forced diuresis, the patient was discharged with proximal paraparesis to rehabilitation with a diagnosis of acute myopathy.

From discharge to 3-month follow-up, we observed 4 patients with PNS complications. Two ICU patients complained of burning paranesthesia and numbness of the lateral thigh (bilaterally in one case) and were diagnosed as having meralgia paresthetica. Both patients had been ventilated in the prone position leading to compression of the lateral femoral cutaneous nerve.

Two patients developed a peripheral facial palsy after discharge. One patient had a right-sided palsy together with a cutaneous rash in the neck 3 months after a positive SARS-CoV-2 test, and microbiological cultures and antibody tests of the skin lesion confirmed varicella zoster virus infection (VZV). The CSF was notable for a marginal pleocytosis of 7, protein of 0.7g/L, negative borrelia, VZV, herpes simplex virus, enterovirus screening and negative CXCL-13. The second patient developed peripheral facial palsy 40 days after a positive SARS-CoV-2 test; there were <3 cells in the CSF, protein was 0.16g/L, and screening for herpes simplex virus, VZV, Epstein Barr virus and cultures were negative. Neither of the patients’ CSF specimens were screened for SARS-CoV-2.

One patient was seen 5 weeks after discharge with a right-sided foot drop, mild dysphagia and a tongue palsy deviating to the left. Brain MRI was unremarkable, and ENG showed mild bilateral peroneal nerve affection. Symptoms resolved spontaneously after 2 weeks. CSF was not investigated.

We used a logistic regression model with the covariates of total admission days, dialysis, delirium, ICU admission and age, and found that the duration of admission was an independent risk factor for (possible) CIPM with an increase of OR=1.14 (95% CI=1.00-1.32) for each day of admission, p=0.046.

### 3.5 Case definitions

The observed complications as outlined in 3.4 were classified in 4 different groups based on a case definition approach described in Methods:

No complications met criteria for ‘direct viral invasion’ (A), 3 complications met criteria for ‘immune-mediated mechanisms’ (B), 34 complications were categorized as ‘complications secondary to critical illness, management-related or other causes’ (C) and 4 complications which could not be categorized as A, B or C were labeled ‘insufficiently investigated’ (D).

## 4. Discussion

In this prospective study of consecutively enrolled hospitalized COVID-19 patients, half of which were admitted to the ICU, we evaluated the semiology, frequency and cause of CNS and PNS complications of COVID-19 during admission and at discharge, including 3-month follow-up. As hypothesized, these complications were immune-mediated or treatment-related and more frequent and serious in COVID-19 patients requiring ICU management. The most common CNS complication was encephalopathy encountered in roughly ⅓ of all COVID-19 patients, with duration of ICU admission being an independent predictor. The most common PNS complication was CIPM, seen in 13% of patients, again with length of admission as an independent predictor. After applying our case criteria described in Methods, only 3 observed complications met criteria for SARS-CoV-2 associated immune-mediated neurological disease: 2 encephalitides and 1 myelitis. All other complications were classified as either secondary to critical illness or other causes or were insufficiently investigated.

### 4.1 Encephalopathy

Encephalopathy affected roughly a third of COVID-19 patients, which is consistent with a recent retrospective study [19], yet considerably lower than previous reports from the early phase of the pandemic (69%) [1], and much higher than another, prospective recent report (6.8%) [15]. This probably reflects differences in methodology as outlined below.

Interestingly, we encountered 8 patients with akinetic mutism, i.e. they were apathetic and profoundly hypokinetic with almost no voluntary movements and without any verbal output, although they were able to indicate yes and no by nodding with their heads when spoken to. On neurological exam, all these patients had some frontal release signs (e.g. palmomentum reflexes), and during our systematic daily clinical evaluation we found this to be a transient phenomenon lasting for a median of 4 days. Akinetic mutism has been described in a COVID-19 case report, lasting 5 days [20], and in a case-series of 4 patients [21]; all 5 patients had frontal FDG-PET hypometabolism, while CSF and MRI findings were normal [20,21]. Akinetic mutism is typically associated with frontal lobe injury [22], and we indeed observed frontal lobe abnormalities, including a small frontal lobe cortical subarachnoid hemorrhage in one patient and FIRDA in 3 of 4 patients investigated with EEG, while MRI in 3 and CSF in 2 patients were unremarkable. Seven of our 8 patients had long ICU admissions (median 21 days) and sepsis with multiorgan injury, and all had delirium. Indeed, hypokinetic delirium, which has clinical similarities to akinetic mutism, is the most frequent motor subtype of delirium [23], and brain FDG-PET of delirious patients may show cortical hypometabolism, most frequently in the frontal lobes [24].

### 4.2 Encephalitis and myelitis

We observed 3 cases of CNS inflammation compatible with para- or post-infectious immune mediated complications to COVID-19 (**Figure 3**). One patient was diagnosed with probable COVID-19-associated acute necrotizing encephalitis, after extensive laboratory evaluation including a brain biopsy and a whole body FDG-PET ruled out other microbiological, autoimmune or neoplastic causes. Three previous cases of acute necrotizing encephalitis have been reported with COVID-19 [25–27]. All 3 patients, like ours, were females aged 30 to 40 years with neurological symptoms within 2 weeks of COVID-19 onset. Two cases [25,26] were believed to be caused by an indirect immune-mediated hyperinflammation (i.e. a cytokine storm) [28], which also has been reported with other viral infections including influenza [29,30], whereas in the third case [27] low levels of SARS-CoV-2 RNA were detected in the CSF indicating possibly direct viral neuronal injury.

Our second encephalitis patient had negative SARS-CoV-2 PCR but positive SARS-CoV-2 antibody testing in CSF. Given the pleocytosis of 250 cells, a protein of 1.13 g/L and a negative viral PCR, a plausible explanation for the positive antibody test would be blood contamination due to a leaky blood-brain-barrier and not direct viral invasion. Patients with neurological symptoms and positive SARS-CoV-2 PCR in CSF samples have been reported [11,12], but the majority of patients with neurological manifestations investigated with PCR on CSF had undetected levels of SARS-CoV-2 [8]. This was consistent with our finding that all 5 CSF samples investigated for SARS-CoV-2 were PCR negative.

Our third case of probable immune-mediated CNS inflammation was a patient with monophasic myelitis who made an uneventful recovery, with neurological symptom onset within 14 days after COVID-19 symptom debut and negative SARS-CoV-2 CSF PCR. As in another reported case [31], investigations for other causes of myelitis were negative. Long-term follow-up is needed to determine the risk for recurrence.

### 4.3 Neuromuscular complications

Eight of our patients had hypotonic tetraparesis with hypo- or areflexia and muscle atrophy. Unfortunately, owing to lockdown measures to prevent the spread of SARS-CoV-2, only one patient was investigated with nerve conduction studies which were consistent with CIPM. All 8 patients had prolonged admissions to the ICU, sepsis and multi-organ failure, resembling typical ICU-acquired weakness [32]. Previous studies on PNS complications in COVID-19 have mainly focused on Guillain Barré syndrome [33] which we did not observe, even though our center is the referral center for GBS cases in the Greater Copenhagen area with 1.9 million inhabitants. We did, however, observe one case of rhabdomyolysis as a presenting complication of COVID-19, and myalgia with elevated CK in COVID-19 has indeed been previously reported [33]. Our patient with rhabdomyolysis and acute myopathy had no obvious cause for muscle injury other than COVID-19, but we were unable to obtain a muscle biopsy to determine if there were signs of direct viral invasion or inflammatory cell infiltrates.

Two of our patients were diagnosed with meralgia paresthetica, which has been reported as a complication following mechanical ventilation in the prone position in non-COVID-19 [34] and in COVID-19 patients [35]. As prone positioning in acute respiratory distress syndrome (ARDS) is strongly recommended [36] (and was performed in 61.7% of our ICU population), attention to peripheral nerve injury in COVID-19 ICU survivors seems warranted [37].

Further 2 patients developed a peripheral facial palsy after discharge. One patient had a concomitant VZV infection affecting the neck which was felt to be the most probable underlying etiology of the peripheral facial palsy, while the second patient had no plausible cause other than a recent SARS-CoV-2 infection. Unfortunately, the patients’ CSF samples were not tested for SARS-CoV-2 RNA. In a case-series of 8 patients with COVID-19 associated peripheral facial palsy, all CSF studies were without inflammation (as in our cases), and 5 had PCR testing for SARS-CoV-2 in the CSF with negative results in all [38].

### 4.4 Methodological considerations

Besides prospectively screening for CNS and PNS symptoms during admission and at discharge, we adapted a case definition approach [7] to categorize complications into distinctive groups of suspected disease mechanisms. The vast majority (83%) was categorized as ‘complications secondary to critical illness, management-related or other causes’; much fewer complications were categorized as ‘indirect immune-mediated’ (7.3%); and none were categorized as “direct viral invasion”. A case definition approach has the benefit of attributing neurological disease to COVID-19 only when extensive laboratory investigations have been performed and other known causes have been excluded. This is important to avoid falsely assigning symptoms and signs to COVID-19 when in fact there is unrelated pathology: We encountered a remarkable patient who was diagnosed as having cerebral autosomal dominant arteriopathy with subcortical infarcts and leukoencephalopathy (CADASIL) prompted by typical neuroimaging features, including bilateral anterior temporal lobe FLAIR hyperintensities, on MRI performed for prolonged encephalopathy following COVID-19 (online appendix, **Figure S2**). We did not include this patient in our prospective cohort because she presented to us several months after COVID-19. A similar case of CADASIL diagnosed in a COVID-19 patient owing to unsuspected MRI features has recently been described [39].

Our study has several other strengths: We evaluated a complete and consecutive sample of patients from a tertiary care center with detailed clinical characteristics, and we did not restrict inclusion to patients with a priori neurological presentations but enrolled all patients admitted, irrespective of their clinical symptoms. We distinguished between symptoms, investigations and clinical diagnoses. Unlike retrospective studies that depend on correct EHR coding for data extraction [1–4], we examined patients systematically and prospectively during the entire hospital stay and at discharge, thereby reducing the likelihood for underestimation of CNS and PNS complications. Like our study, a recent large prospective study [15] reported on pre-specified diagnoses but, unlike ours, its data was extracted from chart review of notes provided during neurology hospital consultation service. In our experience, not all patients are referred for neurologist consultation but are managed by anesthesiologists and infectious disease specialists instead, which probably leads to care providers overlooking neurological complications to some degree.

As to limitations, our cohort size was small, owing to the comparatively low burden of COVID-19 in the general population in Denmark in early 2020 [40]; patients were highly selected as ours is a tertiary referral center, which limits the generalizability of our findings; not all patients were sufficiently investigated; and we did not include a control group of non-COVID-19 patients. Future case-control studies are necessary to determine if SARS-CoV-2 infections are causal or coincidental and if complications are indeed more prevalent in COVID-19 patients compared to non-COVID-19 patients.

## 5. Conclusions

Neurological complications are common in hospitalized COVID-19 patients, occur mainly in severely affected patients in the ICU and can often be attributed to critical illness. In cases in which neurological disease was likely directly associated with COVID-19, we did not find signs of viral invasion of SARS-CoV-2 into the CNS or PNS, but laboratory inflammatory changes suggested autoimmune-mediated para- and post-infectious mechanisms. In this consecutive sample of 61 COVID-19 patients admitted to a tertiary hospital, the most common CNS and PNS complications were encephalopathy and CIPM, respectively, while other specific neurological presentations included akinetic mutism, acute necrotizing encephalopathy, transverse myelitis (autoimmune-mediated, CNS), and meralgia paresthetica (treatment-related, PNS).

## Data Availability

De-identified data can be made available to qualified investigators upon reasonable written request to the corresponding author.

## Notes

**Funding:** This work was supported by the Lundbeck Foundation (R349-2020-658).

### Competing Interest Statement

The authors have declared no competing interest.

### Funding Statement

This work was supported by the Lundbeck Foundation (R349-2020-658). The sponsor had no role in the acquisition of the data, interpretation of the results or the decision to publish the findings. Conflict of Interest Disclosures: The authors have no relevant financial disclosures

