## Supplemental files for "Central and peripheral nervous system complications of COVID-19: A prospective tertiary center cohort with 3-month follow-up"

F**igure S2**

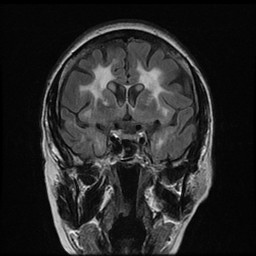

**A1**

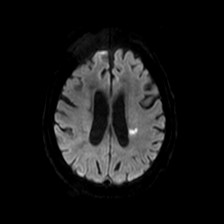

**B2**

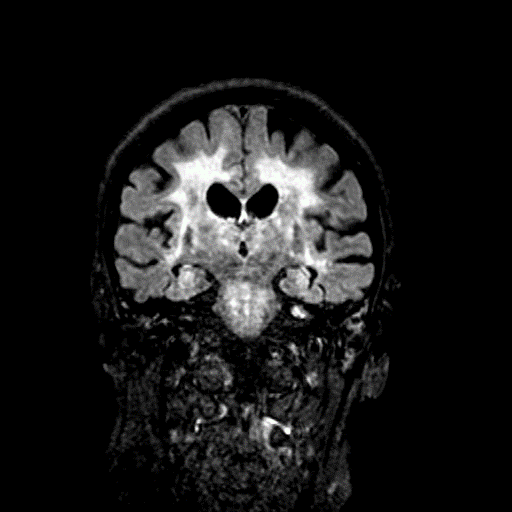

**B1**

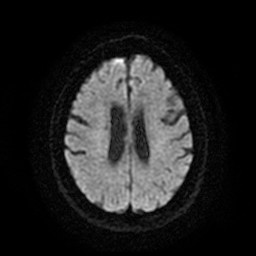

**A2**

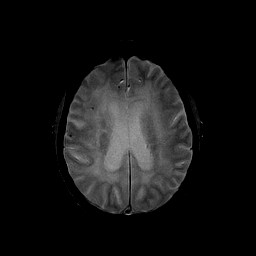

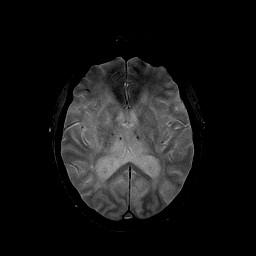

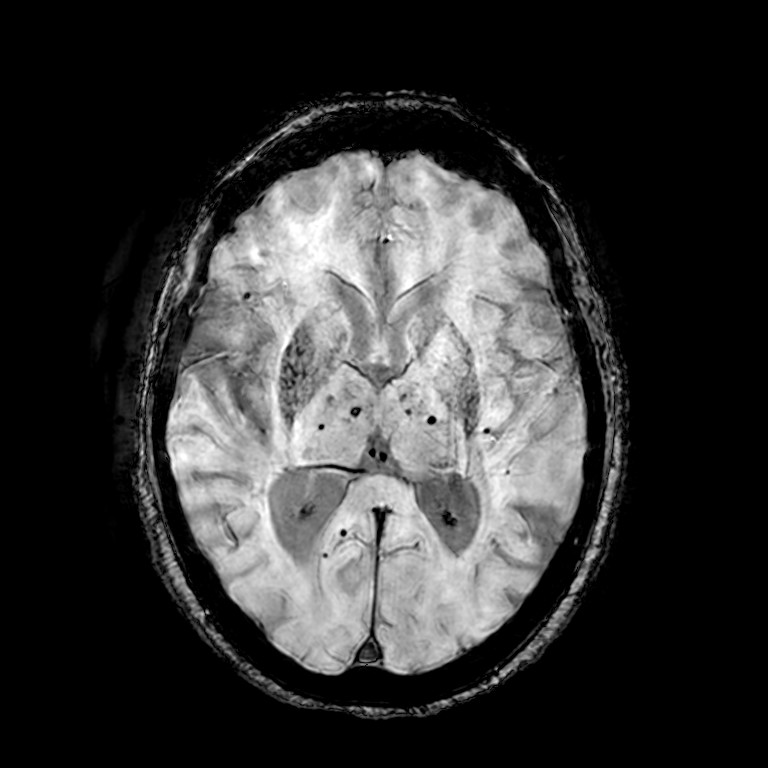

**B3**

**A3**

**A4**

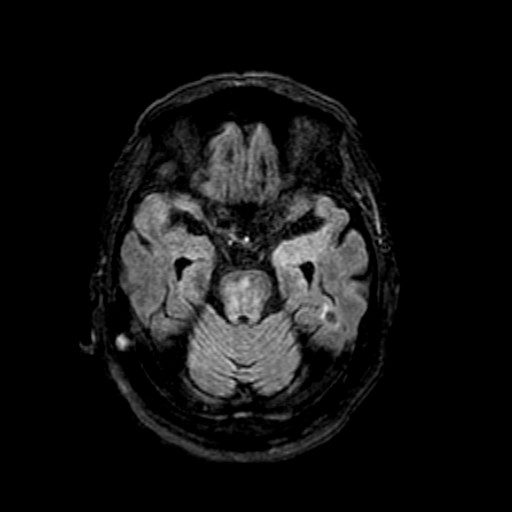

**B4**

**Figure S2:**
A 70+-year-old patient was admitted due to 5 days of fever, malaise and progressive respiratory insufficiency, with a positive SARS-CoV-2 PCR test on pharyngeal swab. She had a previous medical history of hypertension, hypothyroidism, previous cerebellar ischemic stroke and recurrent falls that had been investigated a year prior to admission (A1-A4) at an outpatient geriatric clinic, with magnetic resonance imaging (MRI) of the brain, coronal FLAIR (A1) showing severe leukaraiosis (Fazekas grade 3) and anterior temporal lobe hyperintensity (arrow), but no hyperintense lesions on diffusion-weighted imaging (DWI) (A2). T2*-weighted imaging showed few microbleeds in the basal ganglia (A3) and centrum semiovale (A4). Prior to her COVID-19 admission there was a history of mild cognitive decline, but otherwise the patient had an independent function of daily activities without need for nursing care (Modified Rankin Scale, mRS = 1).
The patient was admitted for a total of 40 days to the intensive care unit (ICU) for her COVID-19 induced respiratory failure. After sedation stop there was a prolonged awakening phase and several days without any verbal output or spontaneous movements, although the patient could follow the examinator with her eyes. There were no focal deficits, but a general ICU-acquired weakness and cognitive deficits interpreted as delirium. After neurological consultation, an MRI scan of the brain was performed that showed (B1) severe leukaraiosis (Fazekas grade 3) and cortical atrophy which was assessed as being more severely than the MRI a year prior (A1). DWI showed a hyperintense signal in the left corona radiata (B2) and susceptibility-weighted imaging (SWI) showed microbleeds in the basal ganglia (B3) and axial FLAIR showed anterior temporal pole hyperintensity (B4, arrow). Initially, a suspicion of cerebral amyloid angiopathy and a COVID-19 associated thrombotic disposition was believed to be the cause of the MRI findings, and the patient was transferred to the neurological department for rehabilitation. An extended family history was taken and revealed several cases of cognitive decline in an early age and together with the patients progressive small-vessel disease, a suspicion was raised for cerebral autosomal dominant arteriopathy with subcortical infarcts and leukoencephalopathy (CADASIL). Genetic testing was performed and reveled a known pathogenic heterozygote mutation in exon 1-25 of the NOTCH3 gene.
